# Patterns of Influenza Vaccine Uptake by Age Group and Product Type in Michigan, 2017-2021

**DOI:** 10.64898/2026.10.19.26365920

**Authors:** Zirui Zhou, Ryan Malosh, Sukhesh Sudan, Abram Wagner, Marisa C. Eisenberg, Arnold S. Monto, Emily T. Martin

## Abstract

**Background:** Influenza vaccines have seen many changes over recent years in terms of both policy recommendations and product types. Immune responses and vaccine effectiveness vary by vaccine strain composition and product type. This study aimed to explore this variation on a regional level through examining spatial and temporal autocorrelations of vaccine uptake of different product types within Michigan from 2017 to 2021.

**Methods:** Influenza vaccine administration data from the 2017/18 to 2021/22 seasons were obtained from the Michigan Care Improvement Registry. Population size estimates for 2017 and 2019 were obtained from the Michigan Department of Health and Human Services. Age-standardized and age-stratified vaccination rates were computed. Disparate county-level vaccine uptake clusters were constructed using spatial autocorrelation indicators and spatiotemporal clusters were identified using scan statistics. Linear mixed effect models were used to study the impact of social vulnerability and urbanicity level on seasonal influenza vaccination rate.

**Results:** Vaccination incidence rates for adjuvanted, recombinant, and nasal sprays vaccines and for any vaccines increased significantly through the study period. County-level spatiotemporal clusters of type-specific uptake rates were identified. Most notably, we identified an emerging cluster with high uptake of adjuvanted vaccine radiating from Ottawa County, located in suburban Grand Rapids, and the disappearance of high uptake cluster for high-dose vaccine uptake in the area. Vaccination rates by any product type were found to vary seasonally and showed weak negative association with more rural regions. The overall vaccination rates appear to be lower among counties with higher social vulnerability in certain urbanicity levels, but social vulnerability and urbanicity were not significantly associated with overall vaccination rate during the study period.

**Conclusions:** Vaccine type-specific uptake was spatially and temporally diverse in Michigan between 2017 and 2022. Mid and Northern Michigan had consistently relatively lower vaccination rate compared to other regions in Michigan.

## Introduction

Influenza virus is a segmented and enveloped RNA virus capable of rapid evolution through antigenic drift and shift resulting in annually antigenically distinct strains [1], which has been posing major infection burdens [2]. Seasonal vaccination remains the most effective preventative measure against influenza infection and hospitalization [3]. Three product types of influenza vaccine are currently in use with variations, including egg- and cell-based inactivated influenza vaccine (IIV), live attenuated influenza vaccine (LAIV), and recombinant influenza vaccine (RIV) with comparable vaccine effectiveness across populations and limited evidence showing better protection in the elderly from RIV [4,5]. Enhanced products designed for high-risk older population including high dose and adjuvanted vaccines have been rolled out in the past decade, providing evidently improved protection against infection [6,7]. Despite continuous effort in vaccine development, influenza infections remain a challenge for preventative measures. Antigenic drift between vaccine strains and circulating strains propagation pathway during vaccine production [8] and strain selection mismatch [9] have been suspected to be contributing factors for the low and varied effectiveness across seasons. Influenza vaccine production method leads to vaccine components and delivery methods triggering different immune responses and the effectiveness of different influenza vaccine product types has been extensively studied to vary on the populational level [4–6,10,11]. Therefore, being vaccinated for influenza must be considered in light of the specific product type to understand the corresponding protective effect.

Vaccination policies guide access for different age groups to different products such as high-dose and adjuvanted IIV for elderly population, which also leads to regional difference in product specific uptake [12]. Vaccine cost and insurance coverage along with other socioeconomic factors also heavily impact vaccine uptake [13,14]. Regional vaccine uptake also varies by socioeconomic and urbanization levels [15–17] and has been shown to vary on both state- and county-level in the US for influenza vaccines [18,19]. Influenza vaccine efficacy is known to differ by age group due to the difference in immune history shaped by past vaccination, exposure, and infection history [20]. Similarly, the baseline population immunity against influenza would also vary by regional exposure to infection and vaccine product. The complex interaction between infection and vaccination and life-course accumulation of heterogeneous immune responses present the challenging problem of quantifying the susceptibility of regional due to populations against influenza.

To quantify sub-population susceptibility, understanding the access to vaccine products is essential. However, few studies have examined county-level vaccine uptake by product type for different age groups across multiple influenza seasons. Hence, we aim to examine regional demographic group specific access to vaccine products by analyzing spatiotemporal autocorrelation of influenza vaccine uptake by age group and product type and estimating the relationship of vaccine uptake with social vulnerability and urbanization levels within Michigan, US from 2017 to 2022.

## Methods

### Data sources

We obtained aggregated vaccine administration data based on age group and vaccine product type for five influenza seasons (2017/18 to 2021/22) from the Michigan Care Improvement Registry (MCIR) [21]. MCIR is a life-course immunization information system, with mandatory reporting of all immunizations administered to individuals < 20 years of age in Michigan. Total of nine categories of vaccine product types were included: cell, recombinant, adjuvanted, trivalent egg, quadrivalent egg, high dose, live attenuated, nasal spray, and historic (unspecific vaccine type on record) vaccines. Vaccination records for products administered to individuals in age groups not recommended by CDC were removed from spatiotemporal autocorrelation analysis. County-level population estimates by age group (6 months through 4 years, 5 through 12 years, 13 through 17 years, 18 through 24 years, 25 through 49 years, 50 through 64 years, 65 years and older) for 2017 and 2019 were obtained from Michigan Department of Human Health and Services. We used the most recent population estimates for each influenza season.

County level demographic factors related to vaccine access were downloaded from government data portals. The Social Vulnerability Index (SVI) was created using CDC/ATSDR’s Geospatial Research, Analysis & Services Program to establish an index for comparing the relative vulnerability across communities different regions [22]. We used the overall sum of fifteen county-level variable percentiles from the Socioeconomic, Household Composition & Disability, Minority Status & Language, and Housing Type & Transportation themes of 2016, 2018, and 2020 SVI datasets for Michigan to create a categorical variable representing the four quartiles of the sum (Q1, Q2, Q3, and Q4), with higher quartile indicating higher relative overall social vulnerability for a specific county during the corresponding year [15]. Urbanization levels for each county were defined by the 2013 National Center for Health Statistics Urban-Rural Classification Scheme for Counties with six levels from most urban to most rural: large central metro, large fringe metro, medium metro, small metro, micropolitan, and noncore [23]. Urbanization scores ranging from 1 to 6 were used to reference the 6 metropolitan categories respectively.

### Vaccination rate and Age standardization

We calculated overall and product-specific vaccination rate within each age group for each county as the number of vaccine administration per 100,000 person-years. To check vaccination rate variations by age, we compared vaccination rate ratios across different age groups using 6 months through 4 years old as reference group and calculated the standard error of each ratio for each season [24]. To obtain comparable vaccination rates by product type regardless of different county age distributions, direct age standardization to the state age group structure was applied to age group vaccination rate for each county [25]. To understand vaccination rate’s relationship with accessibility, we examined the association of social vulnerability and urbanicity level with age-standardized any-type vaccination rate using linear mixed effect model incorporating random intercepts for each county. Model selection was based on Akaike’s Information Criterion (AIC) and Bayesian Information Criterion (BIC) for fitness evaluation [26]. Model assumptions were checked to ensure no major violations.

### Spatial and temporal autocorrelation analysis

Spatial weights were created under the Queen’s first order contiguity rule where surrounding counties sharing borders with a county were considered as its neighbors. Spatial clusters based on vaccination rate were constructed using Local Moran’s I and state-level clustering was assess using Global Moran’s I with *rgeoda* package [27]. Local and Global Moran’s I measures the spatial autocorrelation of vaccine uptake for counties and Michigan state respectively in this setting, with positive Moran’s I values indicating clustering and negative suggesting dispersion of similar uptake levels [28]. A county was labeled as having high vaccine uptake status if its vaccination rate was above the mean county-level incidence and was labeled as low otherwise. The mean vaccination rate for all neighboring counties of that county was used to determine the neighbor vaccine uptake status. Four different county-level cluster labels were created using a county’s vaccine uptake status followed up with their neighbor uptake status: High-High, High-Low, Low-High, and Low-Low, describing a county’s and its surrounding counties’ vaccination rates compared to global mean in that order. The significance for Local and Global Moran’s I were determined by comparing observed county vaccination rate to simulated distributions through 999 permutations that randomized county assignments of observed incidence, with p-values < 0.05 being considered as significant and the null value of 0 being tested for both. Spatial-temporal clusters with maximum likelihood estimate and significance level obtained through Monte Carlo simulations assuming no clustering as null were detected using space-time scans statistic using a discrete Poisson mode adjusting for age groups in *SaTScan* software where a cluster’s high or low uptake status was determined by comparing observed to expected vaccine administration counts adjusting for population estimates [29,30]. Specific model parameter settings can be found in the supplemental materials.

All analyses were performed in R 4.4.1 unless specified otherwise.

## Results

### Vaccine uptake across different influenza seasons

We observed negligible changes in age group distribution of Michigan populations between 2017 and 2019 (Table 1). Specific county level population estimates are listed in Supplemental Table 1. Age-group vaccination rate ratios are presented in Supplemental Table 2. Over influenza seasons 2017/18 to 2021/22, the 2020/21 season recorded the highest vaccination rate of 60,762 per 100,000 person-years by any vaccine product types with the 2017/18 season having the lowest vaccination rate of 48,199 per 100,000 person-years. The vaccination rates appeared to vary by age group over the five seasons with two age groups, 65 years and older and 6 months to 4 years, having noticeably higher vaccination rate than other groups (Figure 1). The incidence for 6 months to 4 years old showed a drastic increase from 40,222 per 100,000 person-years in 2018/19 to 71,059 per 100,000 person-years in 2019/20 and experienced a sharp drop to 56,539 per 100,000 person-years by the end of 2021/22 season. The sudden surge and drop of vaccination rate for those 6 months through 4 years old during the study period was due to no immediate eligibility of COVID-19 vaccines for this age group and concerted efforts advocating for influenza vaccination in preparation of potential twindemics. Overall, in concordance with the any-type incidence, the vaccination rates showed gradual increase till the 2020/21 season and decrease over 2021/22 season.

**Table 1.**
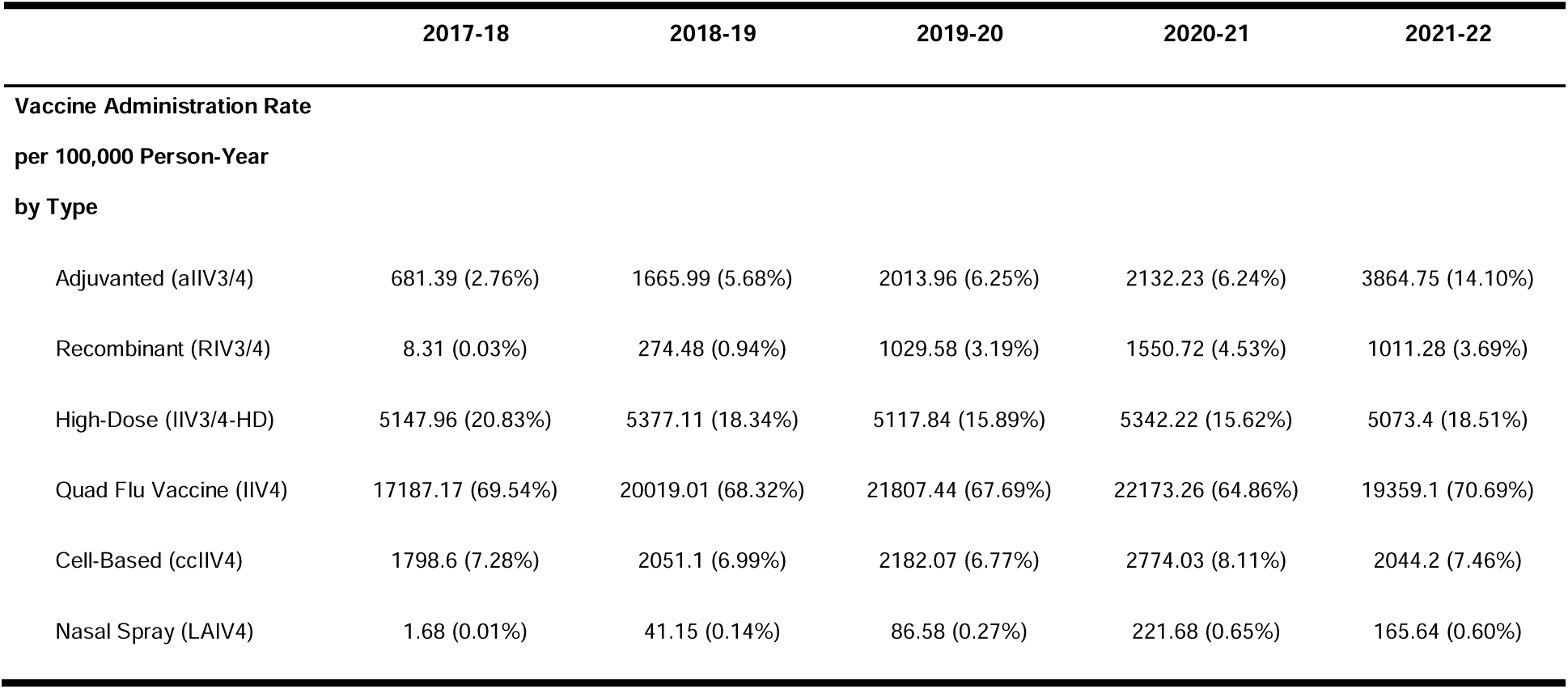
population estimate and age standardized vaccination rate.

### Social vulnerabilities and urbanization levels

The counties in Michigan became less socially vulnerable per CDC/ATSDR’s definition over the study period with 19.3% counties being Q1 in 2016 and 30.1% Q1 in 2020 (Supplemental Table 3). We categorized the counties as 2 large central metro, 8 large fringe metro, 8 medium metro, 8 small metro, 25 micropolitan, and 32 noncore counties. The overall vaccination rates appear to be lower among more socially vulnerable populations with no observable differences by urbanicity levels (Figure 2). The vaccination rate decreased over the study seasons besides an increase in 2020/21 season compared to 2019/20 season and the estimated uptake was generally lower in more rural areas. We observed lower vaccination rate with lower urbanicity level, but this is not statistically significant when controlling for SVI levels (p=0.14).

### Clustering of vaccination receipt

We found areas with disparities in overall vaccine uptake defined as significant high and low clusters based on mean vaccination rates and spatial autocorrelation over 2017/18 to 2021/22 seasons (Figure 3). Based on vaccine incidence alone, Marquette County in Upper Peninsula (UP), Allegan and Barry in Southwest Michigan, and Washtenaw in Southeast Michigan had consistently high incidence above 35000 per 100,000 person-years. Two statistically significant clusters of counties with high vaccine uptake themselves and high uptake neighbors (High-High) were detected radiating out of Allegan County in Southwest and Benzie County in Northern Michigan during the five seasons. Low-low cluster consisting of Macomb, Saint Clair, Lapeer, Sanilac, and Tuscola County in Mid-Michigan was detected since 2017/18 season and began decreasing in size from 2020/21 season. And in 2021/22 season, only Saint Clair County remained in the low-low cluster. Meanwhile, another low-low cluster spreading out from Iosco County in Northern Michigan was detected during the 2019/20 season and formed a larger low-low cluster with five counties at the end of 2021/22 season. The state-wide spatial autocorrelation was also measured to have positive Global Moran’s I from 2017/18 to 2021/22, which implied state-level clustering of counties with similar uptake status and indicated increasingly regionally disparate vaccine uptake within Michigan.

Distinctive spatiotemporal clusters of high and low uptake regions for recombinant and enhanced vaccines (including recombinant, high dose, and adjuvanted products) targeting high risk populations were identified. Low recombinant uptake cluster consisting of 49 counties in Upper Peninsula, Northern, and Southwest Michigan existed from 2017/18 to 2018/19 season. Three high recombinant uptake clusters were founded consisting mostly of Southeast Michigan counties over 2019/20 to 2021/22 seasons (Figure 4a). For adjuvanted vaccines, two low uptake clusters consisting of four and five counties respectively along the southeast state boarder were detected during 2017/18 and 2018/19 seasons (Figure 4b). There was also an emerging high adjuvanted vaccine uptake cluster of 24 counties radiating northeastward from Ottawa County in 2021/22 season. The high dose vaccine had a high uptake cluster with fourteen counties spanning northward from Ottawa County from 2017/18 to 2018/19 season (Figure 4c). This high uptake cluster disappeared after 2018/19 and shifted to Southeast Michigan and reduced noticeably in size with a cluster with two counties in 2018/19 and another with one county during 2020/21 and 2021/22 seasons. Furthermore, one low high dose uptake cluster of six counties was detected in Mid Michigan from 2018/19 to 2019/20 and another one with two counties was seen in Southeast Michigan during 2019/20 and 2020/21 seasons. Given the focus on adults 65 years and older for adjuvanted and high dose products, a sensitivity analysis was performed to view age-specific vaccination uptake (Supplemental Figure 1). Age-specific maps were found to show comparable results to age-standardize patterns, regardless of the presence of vaccine recommendations for targeted age groups over time.

## Discussions

### Identification of low uptake regions

We have identified areas within Michigan that have low uptake for specific types of vaccine between 2017 and 2022. Mid and Northern Michigan were two regions with consistent low vaccine uptake for most vaccine types and more programs promoting vaccination awareness and availability might be needed in those two regions. Vaccine uptake for each vaccine type was also spatially diverse in Michigan and such impact on population level immune response should be studied. Especially for those aged 65 years and above at higher risk of severe symptoms due to influenza infections, there were strong spatial and temporal variability for the uptake of two vaccines (adjuvanted and high dose vaccines) recommended for this age group.

### Spatiotemporal and demographic uptake variations

We found strong vaccine uptake spatiotemporal variability within any specific vaccine type and across different types in Michigan from 2017 to 2022. Despite the increasing vaccination rate by any product type in vulnerable groups, regions with varied product-specific uptake were characterized. Most intriguingly, the adjuvanted and high dose spatial-temporal clusters suggested high dose vaccine was the more prevalent type of the two in Southwest Michigan until 2018/19 season, and high dose overtook it as the more prevalent vaccine in the region after then. From clustering of recombinant vaccine uptake, the roll out sequence of the vaccine could be observed as the vaccine was more widely introduced in Southeast Michigan and consistently had higher uptake in that region than rest of the state. For the 2021/22 season, a general decrease in influenza vaccination rate was observed across all age groups in conjunction with the beginning of the COVID-19 pandemic.

Some regions with both higher social vulnerability and lower urbanicity level were observed to have lower age-standardized vaccination rates, which is consistent with previous findings [31,32], despite such relationship being statistically insignificant. After adjusting for social vulnerability and urbanization, season remained as a good estimator for vaccination rate, suggesting some other factors were promoting vaccination rate and temporal changes of vaccine uptake within Michigan during the study period.

### Implication for population immunity

The spatiotemporal differences in vaccine product uptake complicate efforts to use vaccination patterns to predict population level protection against influenza infection or population immunity, especially in a local setting. Vaccine production method matters since the resulting antigen contained would be different, leading to different mechanisms through which the vaccines elicit immune responses. IIV contains inactivated influenza viruses and is administered through injection while LAIV is given as a nasal spray with weakened viruses. These two types of vaccines are produced using influenza viruses grown in chicken eggs, and more recent cell-based production method has made cell-based IIV available. Influenza viruses grown in eggs are known to acquire adaptive mutations that lead to differing effectiveness for egg-based IIV compared to the cell-based counterpart [33]. RIV is also cell-based and only contains surface protein hemagglutinin, which does not induce neuraminidase specific antibody production, compared to whole viruses in IIV and LAIV. For enhanced vaccines, high dose contains four times the normal amount of antigen and adjuvanted uses the normal amount in combination with adjuvants stimulating immune responses. The effectiveness of different influenza vaccine product types has been extensively studied to vary on the populational level with egg adaptation and strain mismatch between vaccine and circulating strains being speculated as major contributors to vaccine effectiveness variations [4–6,10,11]. To the degree that different vaccine products result in different magnitude and breadth of immune response, differences in regional availability and uptake of enhanced vaccine product types could potentially result in regional differences in the magnitude and breadth of protection against current and emerging strains. Hence, composing population immunity profiles accurately would most likely require knowledge of regional vaccine uptake by product type on top of other factors including as age structure and birth cohort due to immune imprinting by circulating strains during early childhood [34,35].

### Limitation

Although Moran’s I is a well-established indicator of local indicator of spatial association [36], we did not explore whether the clustering results stayed consistent when using other clustering metrics such as isolation index and Theil index that had been used in other similar studies [37]. Data artifacts might be present in vaccination record used in this study. There were records for vaccination of types that were not recommended for specific age groups such as high dose vaccinations among 6-months-through-4-year-olds. The quality of MCIR vaccination registry records have been validated but could be under-reporting vaccinations among adults whose records are not required to be reported unlike children’s and from providers that do not report using records captured by MCIR [38]. The spatial resolution of this project is set to county level but can be further aggregated to health department district level to examine the impact of local vaccination policy changes in Michigan. Future research might also want to explore vaccine uptake on a higher spatial resolution such as census tract level to examine effects of differing vaccination campaigns in neighboring areas.

## Conclusion

This study provides a detailed snapshot of changes in influenza vaccination uptake through the initial years of the COVID-19 pandemic, a tumultuous time for vaccination communications and policy. Given the ongoing uncertainty in vaccine recommendations, this work provides a blueprint for the level of detailed monitoring of patterns that is needed. By understanding vaccine use by product, age, urbanicity and SVI, we will be better poised to anticipate the impact of changing vaccine use on population-level protection against influenza, informing more accurate forecasting efforts.

## Supporting information

Supplemental Table 1. Michigan population estimates

Supplemental Table 2. vaccination rate ratio with standard error

Supplemental Table 3. SVI distribution by Michigan Counties

Supplementary Table 4. Final Mixed-Effects Regression Model Results

Supplemental Figure 1. Sensitivity analysis for Age65+ vaccine uptake trend

## Data Availability

All data produced in the present study are available upon reasonable request to the authors

## Acknowledgement

The authors would like to thank Sarah Cobey, Marcos Costa Vieira, and Manon Ragonnet for feedback on the analysis pipeline and appropriate conclusions.

## Disclaimer

This work was made possible by cooperative agreement CDC-RFA-FT-23-0069 from the CDC’s Center for Forecasting and Outbreak Analytics. Its contents are solely the responsibility of the authors and do not necessarily represent the official views of the Centers for Disease Control and Prevention.

---

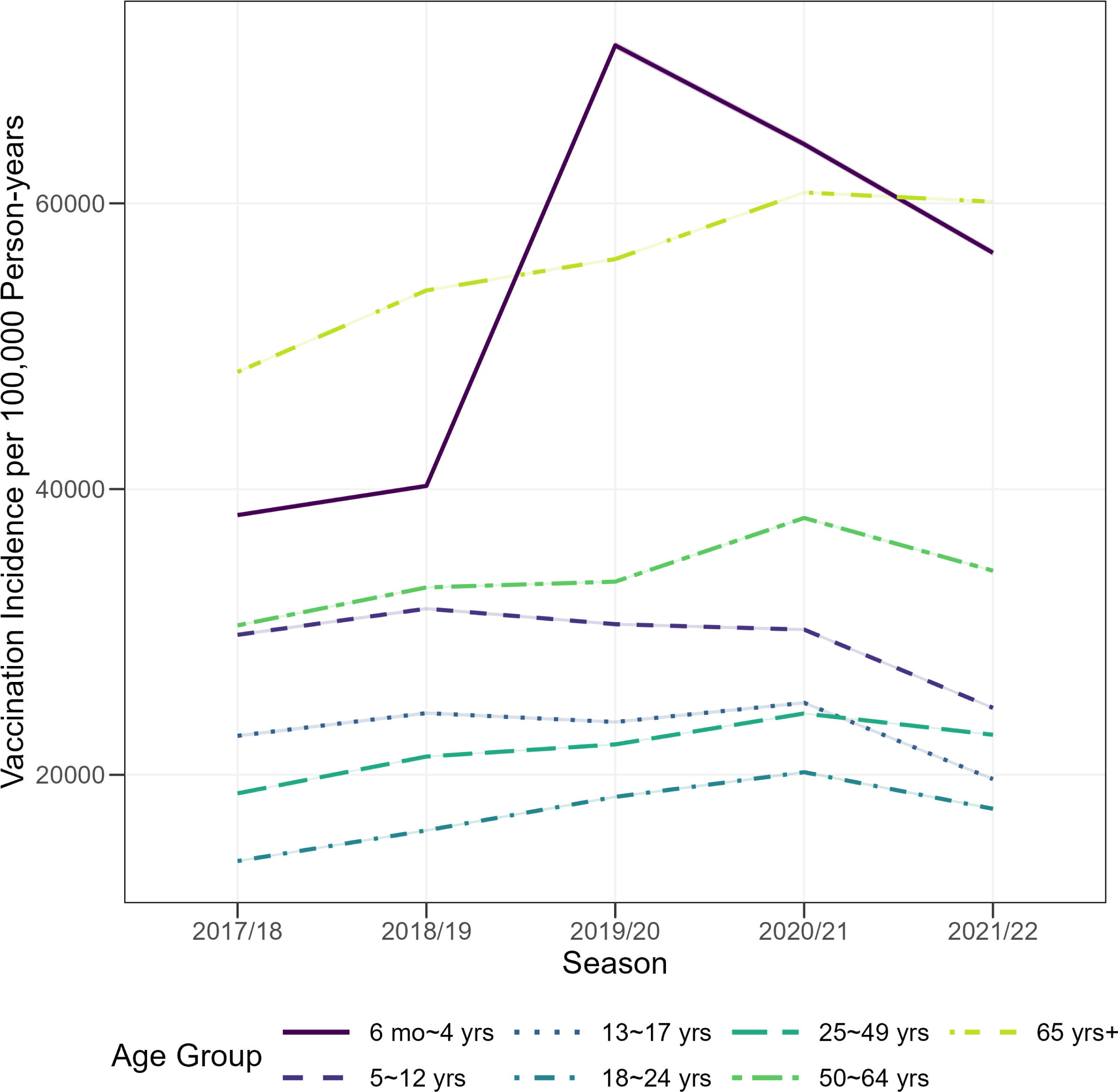

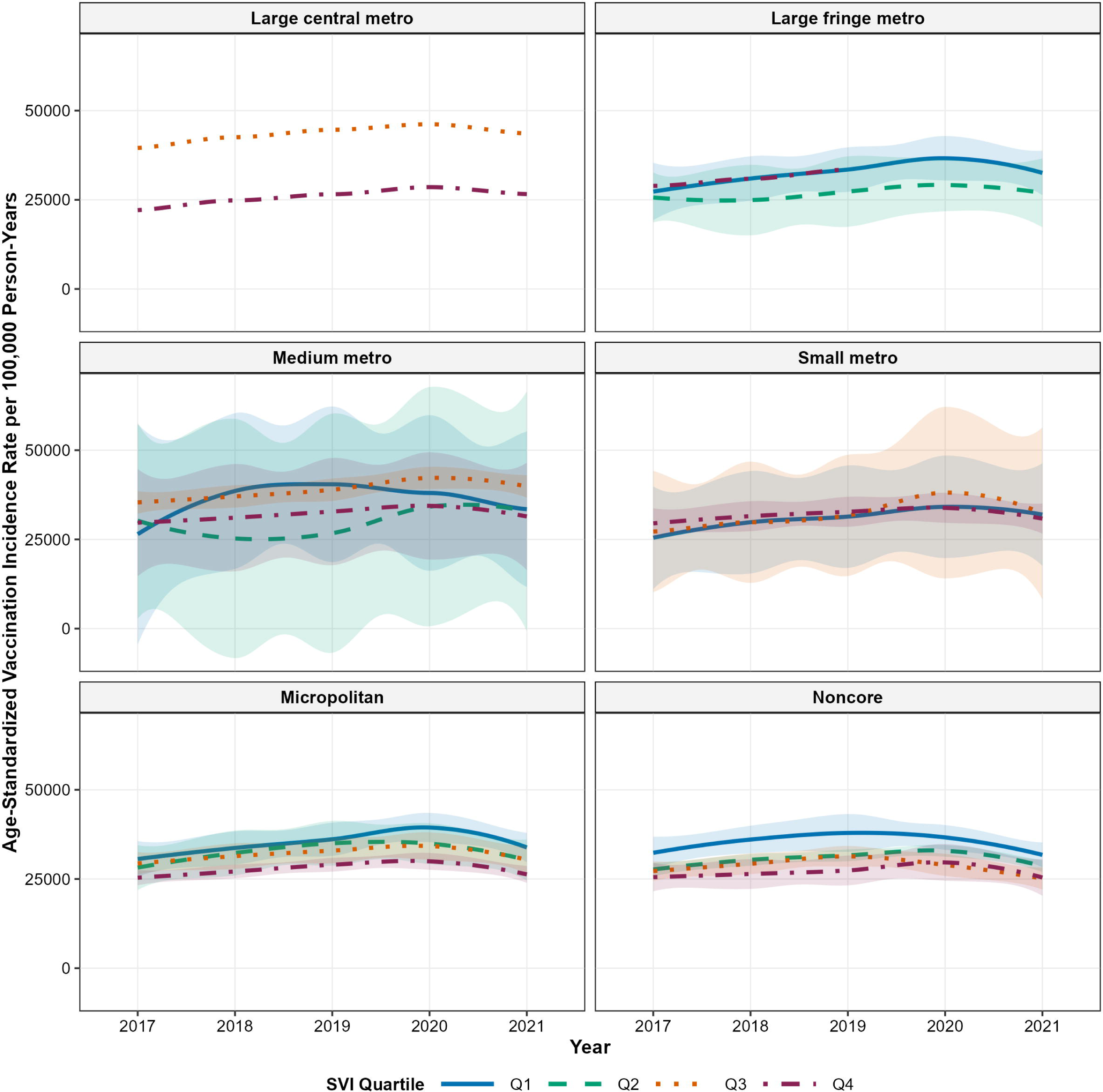

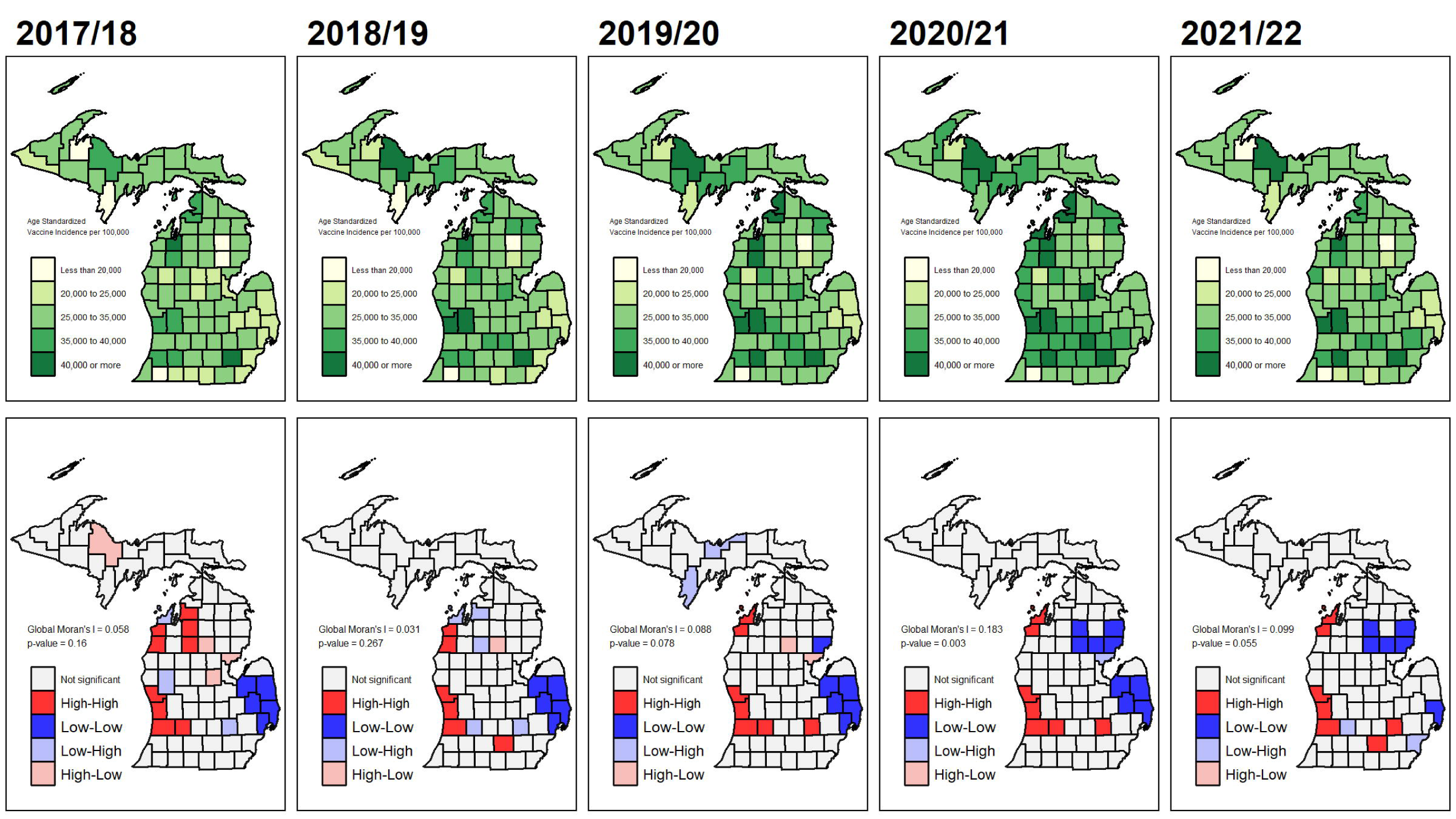

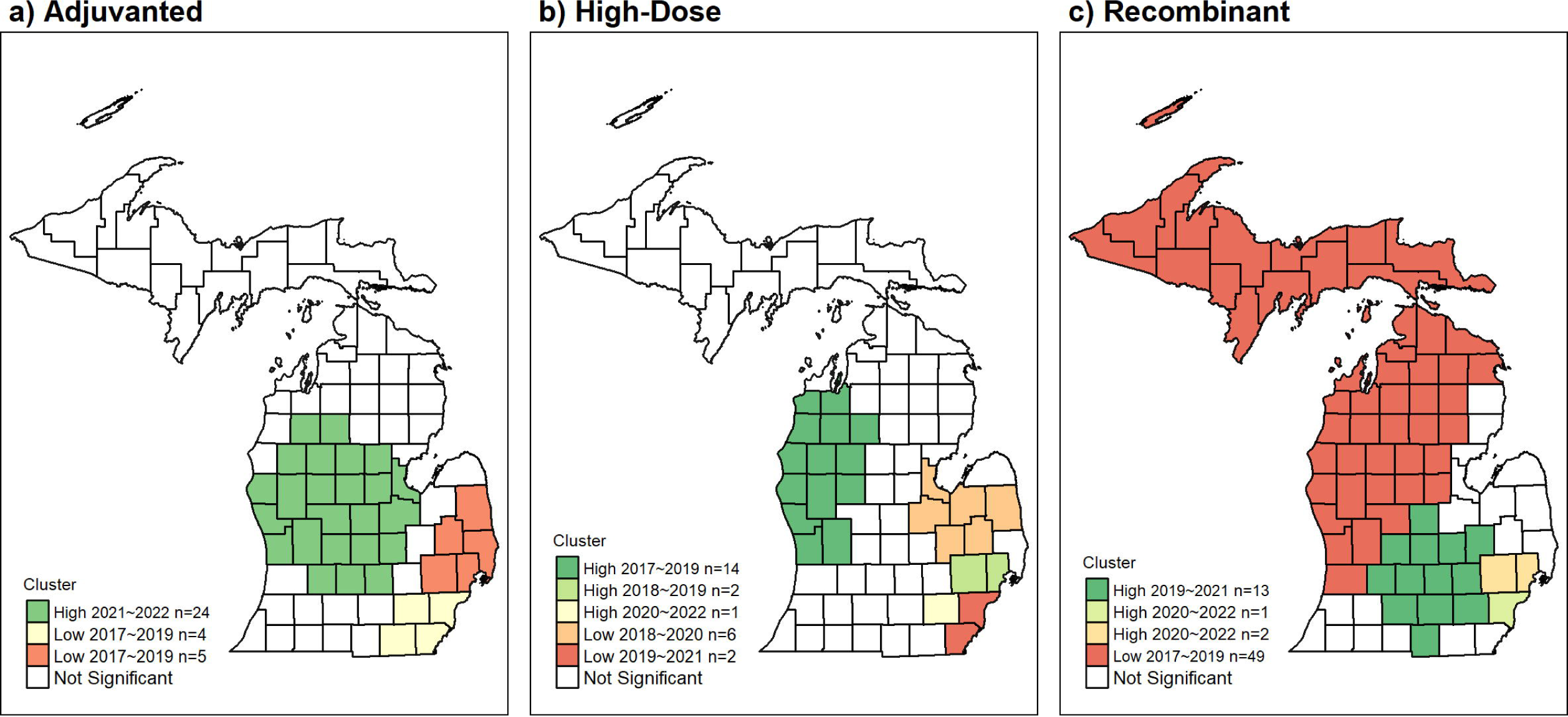

