## Supplemental Table 1. Michigan population estimates for "Patterns of Influenza Vaccine Uptake by Age Group and Product Type in Michigan, 2017-2021"

|  | **2017-18** | **2019-20** |
| --- | --- | --- |
| **Population**  **by Age Group** |  |  |
| 6 months through 4 yrs | 573282 (5.75%) | 404289 (4.03%) |
| 5 through 12 years | 957739 (9.61%) | 1044069 (10.41%) |
| 13 through 17 years | 645628 (6.48%) | 734115 (7.32%) |
| 18 through 24 years | 970807 (9.74%) | 945523 (9.43%) |
| 25 through 49 years | 3067091 (30.79%) | 3090317 (30.82%) |
| 50 through 64 years | 2080568 (20.88%) | 2041683 (20.37%) |
| 65 years and older | 1667196 (16.74%) | 1765401 (17.61%) |
