## Supplemental Table 2. vaccination rate ratio with standard error for "Patterns of Influenza Vaccine Uptake by Age Group and Product Type in Michigan, 2017-2021"

| **Age Group** | **aIIV3/4** | **IIV3/4-HD** | **IIV4** | **ccIIV4** | **RIV3/4** | **LAIV** | **Unspec.** |
| --- | --- | --- | --- | --- | --- | --- | --- |
| **2017~18** |  |  |  |  |  |  |  |
| 6 mo~4 yrs | ref | ref | ref | ref | ref | ref | ref |
| 5~12 yrs | 3.358 (0.452) | 0.485 (0.442) | -0.311 (0.003) | 2.083 (0.037) | -0.395 (0.486) | 0.275 (0.539) | 0.763 (0.020) |
| 13~17 yrs | 3.633 (0.452) | 1.303 (0.421) | -0.633 (0.004) | 2.236 (0.037) | 0.692 (0.425) | -0.630 (0.730) | 0.975 (0.020) |
| 18~24 yrs | 4.247 (0.449) | 2.071 (0.392) | -1.286 (0.004) | 2.334 (0.037) | 1.326 (0.380) | 0.061 (0.558) | 1.306 (0.019) |
| 25~49 yrs | 4.194 (0.448) | 2.935 (0.380) | -1.025 (0.003) | 2.594 (0.036) | 2.030 (0.358) | 0.427 (0.474) | 1.761 (0.018) |
| 50~64 yrs | 5.364 (0.448) | 6.202 (0.378) | -0.587 (0.003) | 3.220 (0.036) | 2.234 (0.359) | 0.628 (0.479) | 2.243 (0.018) |
| 65 yrs+ | 8.345 (0.447) | 10.108 (0.378) | -1.362 (0.003) | 2.124 (0.036) | 1.830 (0.363) | 1.482 (0.464) | 1.895 (0.018) |
| **2018~19** |  |  |  |  |  |  |  |
| 6 mo~4 yrs | ref | ref | ref | ref | ref | ref | ref |
| 5~12 yrs | -0.002 (0.730) | 0.148 (0.308) | -0.291 (0.003) | 1.920 (0.029) | 0.180 (0.340) | 0.769 (0.046) | -0.254 (0.022) |
| 13~17 yrs | 0.862 (0.677) | 0.441 (0.313) | -0.576 (0.003) | 1.962 (0.029) | 1.759 (0.298) | -0.161 (0.058) | -0.213 (0.024) |
| 18~24 yrs | 1.840 (0.604) | 1.782 (0.262) | -1.077 (0.003) | 1.985 (0.029) | 4.294 (0.278) | -2.090 (0.097) | 0.298 (0.020) |
| 25~49 yrs | 2.815 (0.581) | 2.468 (0.252) | -0.828 (0.003) | 2.265 (0.028) | 4.907 (0.278) | -2.490 (0.073) | 0.883 (0.017) |
| 50~64 yrs | 5.774 (0.578) | 5.491 (0.250) | -0.425 (0.003) | 2.762 (0.028) | 5.703 (0.277) | -3.250 (0.115) | 1.273 (0.017) |
| 65 yrs+ | 9.830 (0.577) | 9.322 (0.250) | -1.430 (0.003) | 1.476 (0.029) | 3.996 (0.278) | -2.112 (0.079) | 1.335 (0.017) |
| **2019~20** |  |  |  |  |  |  |  |
| 6 mo~4 yrs | ref | ref | ref | ref | ref | ref | ref |
| 5~12 yrs | -0.794 (0.556) | -1.376 (0.332) | -0.895 (0.003) | 3.089 (0.070) | 1.131 (0.354) | 0.540 (0.034) | -0.795 (0.029) |
| 13~17 yrs | 0.105 (0.500) | -1.777 (0.432) | -1.175 (0.003) | 3.352 (0.070) | 2.413 (0.342) | -0.018 (0.038) | -0.822 (0.032) |
| 18~24 yrs | 0.762 (0.447) | 0.745 (0.229) | -1.495 (0.003) | 3.610 (0.069) | 5.057 (0.334) | -1.779 (0.057) | -0.316 (0.027) |
| 25~49 yrs | 1.405 (0.415) | 1.606 (0.211) | -1.334 (0.002) | 3.784 (0.069) | 5.584 (0.333) | -3.516 (0.070) | 0.282 (0.023) |
| 50~64 yrs | 3.148 (0.410) | 3.092 (0.209) | -1.015 (0.002) | 4.281 (0.069) | 7.225 (0.333) | -4.216 (0.115) | 0.736 (0.022) |
| 65 yrs+ | 8.945 (0.408) | 8.532 (0.209) | -1.793 (0.003) | 3.503 (0.069) | 6.178 (0.333) | -3.860 (0.104) | 0.942 (0.022) |
| **2020~21** |  |  |  |  |  |  |  |
| 6 mo~4 yrs | ref | ref | ref | ref | ref | ref | ref |
| 5~12 yrs | -0.364 (0.124) | 0.001 (0.240) | -0.837 (0.003) | 2.801 (0.052) | 0.134 (0.181) | 0.620 (0.022) | -0.679 (0.029) |
| 13~17 yrs | -0.131 (0.127) | -0.239 (0.267) | -1.044 (0.003) | 3.038 (0.052) | 1.348 (0.167) | 0.091 (0.025) | -0.757 (0.032) |
| 18~24 yrs | 1.369 (0.104) | 2.080 (0.210) | -1.353 (0.003) | 3.320 (0.052) | 4.287 (0.157) | -1.582 (0.035) | -0.353 (0.028) |
| 25~49 yrs | 1.546 (0.100) | 2.803 (0.205) | -1.183 (0.002) | 3.470 (0.052) | 4.753 (0.156) | -2.489 (0.032) | 0.166 (0.023) |
| 50~64 yrs | 1.901 (0.100) | 3.513 (0.205) | -0.842 (0.002) | 4.021 (0.052) | 6.085 (0.156) | -4.525 (0.088) | 0.566 (0.023) |
| 65 yrs+ | 6.133 (0.099) | 8.525 (0.204) | -1.609 (0.003) | 2.813 (0.052) | 4.754 (0.156) | -4.120 (0.078) | 2.014 (0.022) |
| **2021~22** |  |  |  |  |  |  |  |
| 6 mo~4 yrs | ref | ref | ref | ref | ref | ref | ref |
| 5~12 yrs | -0.052 (0.133) | -0.857 (0.302) | -0.908 (0.003) | 1.201 (0.031) | 0.413 (0.354) | 0.594 (0.024) | 0.144 (0.030) |
| 13~17 yrs | 0.160 (0.136) | -0.742 (0.321) | -1.150 (0.004) | 1.450 (0.031) | 1.414 (0.337) | -0.107 (0.028) | 0.163 (0.032) |
| 18~24 yrs | 1.113 (0.119) | 1.177 (0.232) | -1.355 (0.003) | 1.918 (0.030) | 5.205 (0.317) | -2.084 (0.046) | 0.758 (0.028) |
| 25~49 yrs | 1.895 (0.113) | 1.828 (0.220) | -1.102 (0.002) | 2.120 (0.029) | 5.672 (0.316) | -3.505 (0.050) | 1.107 (0.027) |
| 50~64 yrs | 2.383 (0.113) | 3.220 (0.219) | -0.776 (0.003) | 2.500 (0.029) | 7.151 (0.316) | -5.448 (0.149) | 1.584 (0.027) |
| 65 yrs+ | 6.987 (0.112) | 8.613 (0.218) | -1.983 (0.003) | 1.295 (0.030) | 5.474 (0.316) | -5.547 (0.168) | 0.830 (0.027) |
