## Supplemental Table 3. SVI distribution by Michigan Counties for "Patterns of Influenza Vaccine Uptake by Age Group and Product Type in Michigan, 2017-2021"

|  | **2016** | **2018** | **2020** |
| --- | --- | --- | --- |
| **SVI Quartile** |  |  |  |
| Q1 | 16 (19.3%) | 19 (22.9%) | 25 (30.1%) |
| Q2 | 20 (24.1%) | 20 (24.1%) | 22 (26.5%) |
| Q3 | 24 (28.9%) | 21 (25.3%) | 19 (22.9%) |
| Q4 | 23 (27.7%) | 23 (27.7%) | 17 (20.5%) |
