## Supplementary Table 4. Final Mixed-Effects Regression Model Results for "Patterns of Influenza Vaccine Uptake by Age Group and Product Type in Michigan, 2017-2021"

| **Predictor** | **Beta** | **95% CI**^1^ | **p-value** |
| --- | --- | --- | --- |
| SVI Quartile |  |  | 0.3 |
| Q1 | — | — |  |
| Q2 | 2,176 | -1,709, 6,060 | 0.3 |
| Q3 | 2,009 | -4,600, 8,619 | 0.6 |
| Q4 | 5,442 | -1,651, 12,536 | 0.13 |
| Year |  |  | **<0.001** |
| 2017 | — | — |  |
| 2018 | 4,542 | 1,330, 7,753 | **0.006** |
| 2019 | 6,916 | 3,705, 10,128 | **<0.001** |
| 2020 | 11,321 | 8,169, 14,473 | **<0.001** |
| 2021 | 7,928 | 4,776, 11,080 | **<0.001** |
| Urbanicity Code (NCHS) | 220 | -952, 1,392 | 0.7 |
| SVI Quartile * Year |  |  | >0.9 |
| Q2 * 2018 | -2,470 | -7,174, 2,234 | 0.3 |
| Q3 * 2018 | -1,378 | -6,198, 3,442 | 0.6 |
| Q4 * 2018 | -2,327 | -6,770, 2,116 | 0.3 |
| Q2 * 2019 | -1,968 | -6,672, 2,736 | 0.4 |
| Q3 * 2019 | -2,039 | -6,859, 2,781 | 0.4 |
| Q4 * 2019 | -2,338 | -6,781, 2,105 | 0.3 |
| Q2 * 2020 | -3,780 | -8,431, 871 | 0.11 |
| Q3 * 2020 | -2,516 | -7,268, 2,236 | 0.3 |
| Q4 * 2020 | -4,897 | -9,532, -261 | **0.039** |
| Q2 * 2021 | -1,337 | -5,988, 3,314 | 0.6 |
| Q3 * 2021 | -1,874 | -6,626, 2,879 | 0.4 |
| Q4 * 2021 | -2,876 | -7,512, 1,759 | 0.2 |
| SVI Quartile * Urbanicity Code (NCHS) |  |  | 0.3 |
| Q2 * Urbanicity Code (NCHS) | -471 | -1,302, 360 | 0.3 |
| Q3 * Urbanicity Code (NCHS) | -370 | -1,647, 906 | 0.6 |
| Q4 * Urbanicity Code (NCHS) | -1,099 | -2,480, 282 | 0.12 |
| Year * Urbanicity Code (NCHS) |  |  | 0.062 |
| 2018 * Urbanicity Code (NCHS) | -319 | -1,028, 391 | 0.4 |
| 2019 * Urbanicity Code (NCHS) | -346 | -1,055, 363 | 0.3 |
| 2020 * Urbanicity Code (NCHS) | -640 | -1,318, 38 | 0.064 |
| 2021 * Urbanicity Code (NCHS) | -943 | -1,621, -266 | **0.007** |
| SVI Quartile * Year * Urbanicity Code (NCHS) |  |  | >0.9 |
| Q2 * 2018 * Urbanicity Code (NCHS) | 471 | -512, 1,454 | 0.3 |
| Q3 * 2018 * Urbanicity Code (NCHS) | 48 | -952, 1,048 | >0.9 |
| Q4 * 2018 * Urbanicity Code (NCHS) | 235 | -708, 1,177 | 0.6 |
| Q2 * 2019 * Urbanicity Code (NCHS) | 278 | -705, 1,261 | 0.6 |
| Q3 * 2019 * Urbanicity Code (NCHS) | 108 | -892, 1,108 | 0.8 |
| Q4 * 2019 * Urbanicity Code (NCHS) | 64 | -879, 1,007 | 0.9 |
| Q2 * 2020 * Urbanicity Code (NCHS) | 235 | -731, 1,202 | 0.6 |
| Q3 * 2020 * Urbanicity Code (NCHS) | -190 | -1,170, 790 | 0.7 |
| Q4 * 2020 * Urbanicity Code (NCHS) | 248 | -724, 1,221 | 0.6 |
| Q2 * 2021 * Urbanicity Code (NCHS) | -40 | -1,007, 927 | >0.9 |
| Q3 * 2021 * Urbanicity Code (NCHS) | -86 | -1,066, 893 | 0.9 |
| Q4 * 2021 * Urbanicity Code (NCHS) | 87 | -885, 1,060 | 0.9 |
| No. Obs. | 415 |  |  |
| AIC | 7,096 |  |  |
| BIC | 7,265 |  |  |
| Log-likelihood | -3,506 |  |  |
| ^1^CI = Confidence Interval | | | |
