## Supplementary figures and images for "Patterns of Influenza Vaccine Uptake by Age Group and Product Type in Michigan, 2017-2021"

### Supplemental Figure 1. Sensitivity analysis for Age65+ vaccine uptake trend

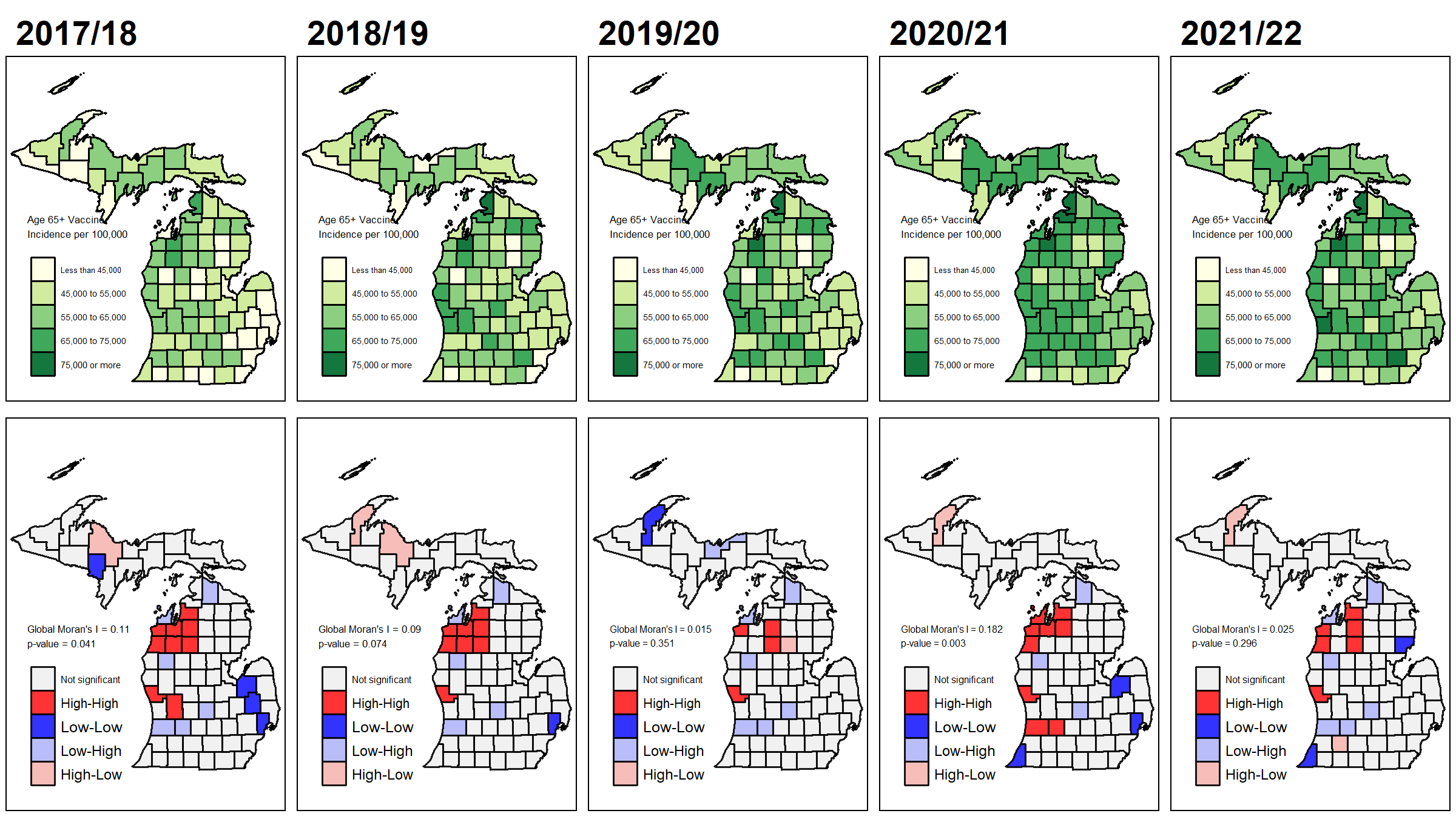
